# A novel nomogram based on serum lipid for identifying the patients at risk for rapid progression of advanced hormone-sensitive prostate cancer

**DOI:** 10.1101/2023.07.07.23292351

**Authors:** Mingshuang Wu, Yi He, Chenxi Pan, Yue Zhang, Bo Yang

## Abstract

**Background:** Serum lipids were reported to be significant predictive factors in various tumors. In order to develop and validate a nomogram for predicting castration-resistant prostate cancer (CRPC) free survival in advanced hormone-sensitive prostate cancer (HSPC) patients, the goal of this study was to assess the prognostic impact of the lipid profiles.

**Material and Methods:** The follow-up information of 146 CRPC patients who received androgen deprivation therapy as the first and only therapy before progression were retrospectively examined. To evaluate prognostic variables, univariate and multivariate Cox regression analyses were used. The concordance index (C-index), calibration curves, receiver operating characteristic (ROC) curves, and decision curve analyses (DCA) were used to design and evaluate a novel nomogram model.

**Results:** Total cholesterol (TC), low-density lipoprotein cholesterol (LDL-C), apolipoprotein B (apoB), N stage and Gleason sum were determined to be independent prognostic markers and were combined to create a nomogram. This nomogram performed well in the customized prediction of CRPC development at 6th, 12th, 18th and 24th month. The C-indexes in training and validation sets were 0.740 and 0.755, respectively. ROC curves, calibration plots, and DCA all suggested favorable discrimination and predictive ability. Besides, the nomogram also performed better predictive ability than N stage and Gleason sum. The Nomogram-related risk score divided the patient population into two groups with significant progression disparities.

**Conclusions:** The established nomogram could aid in identifying the patients at high risk for rapid progression of advanced HSPC, so as to formulate individualized therapeutic regimens and follow-up strategies in time.

## Introduction

With over 350,000 fatalities per year, prostate cancer (PCa) is currently one of the main causes of cancer-specific death in men [1]. The incidence of PCa and advanced PCa have increased since PSA screening was implemented in China, and the rise in cancer-related mortality started in 2012 [2]. By lowering the level of circulating testosterone, androgen deprivation therapy (ADT), a foundation of treatment for individuals with locally or metastatic hormone-sensitive prostate cancer (HSPC), suppresses the progression of PCa. However, the majority of men who receive ADT later on develop castration-resistant prostate cancer (CRPC) within three years [3], with a median survival duration of 14 months [4]. Owing to its heterogeneity, PCa has a broad illness spectrum that includes both aggressive and clinically indolent subtypes. According to Hussain et al’s hypothesis, the risk of death would increase four times if the CRPC stage occurred within the first seven months of ADT [5]. In order to execute an early follow-up plan to rapidly detect progression and optimize treatment regimens, such as chemotherapy, it is vital to clarify the signs that potentially predict progression to CRPC in patients with locally advanced PCa and metastatic PCa [6, 7].

Age, PSA level, Gleason score, tumor stage, lymph node status, and metastasis are several clinicopathological parameters that can offer crucial prognostic information for observing PCa disease progression. However, the predictive ability of these traditional markers is frequently constrained, particularly in patients with a clinical diagnosis that is unclear or who are in intermediate grades or stages.

Lipids are crucial elements of cell membranes and have been linked to the fundamental mechanisms underlying the development of cancer, such as excessive proliferation and abnormal signaling. In addition, cellular energy supply, signaling, and other critical elements of tumor cell proliferation are all influenced by lipid metabolism. Dyslipidemia is frequently noted in individuals with a range of tumor forms because of the accelerated metabolism and proliferation of tumor cells [8]. Recent research has shown that the serum lipid profile is useful in predicting the prognosis of tumors [9, 10], which have been viewed as a possible therapeutic target [11]. Contradictory results were found in the literature on serum lipid levels and PCa. Some results suggested a relationship between them, others refuted this connection [12]. There hasn’t been a definite study on the relationship between lipid markers and CRPC advancement, nevertheless.

In this study, we retrospectively studied the lipid features of patients and investigated their relevance in illness prognosis when paired with current clinical indicators, and further established a lipid profile-based model for predicting the progression of HSPC.

## Methods and materials

### Patient Recruitment and clinico-pathological variables collection

One hundred and forty-six PCa patients with complete follow-up information were participated in this retrospective analysis from November 2016 to July 2022. The protocol was accepted by the ethical committee of the Second Hospital of Dalian Medical University with the Declaration of Helsinki (approval number: 2023064), all patients submitted written informed consent prior to analysis. Clinico-pathological variables at the time of PCa diagnosis, such as age, PSA levels, clinical TNM stage, Gleason sum, total cholesterol (TC), triglycerides (TG), high-density lipoprotein cholesterol (HDL-C), low-density lipoprotein cholesterol (LDL-C), apolipoprotein A1 (apoA1), apolipoprotein B (apoB), and their four derivatives, HDL-C/LDL-C, TG/HDL-C, apoB/apoA1 and HDL-C/TC, were collected and given in Table 1. LHRH agonists (Goserelin 3.6mg specifications: once every 28 days, one dose each time; Leuprorelin 3.75mg specifications: once every 28 days, one dose each time) were injected subcutaneously into various sites on the upper arm, belly, and buttocks for all subjects. combined with anti-androgen (bicalutamide 50mg orally once day or flutamide 250mg orally 3 times daily) as primary and only treatment before advancement. Patients who advanced to CPRC fulfilled the following requirements: 1) serum testosterone level under 50 ng/dl (1.7 nmol/L); 2) PSA progression: PSA level above 2.0 ng/mL, interval 1 week, three times higher than the baseline level > 50 percent. All patients were monitored to CRPC.

**Table 1.**
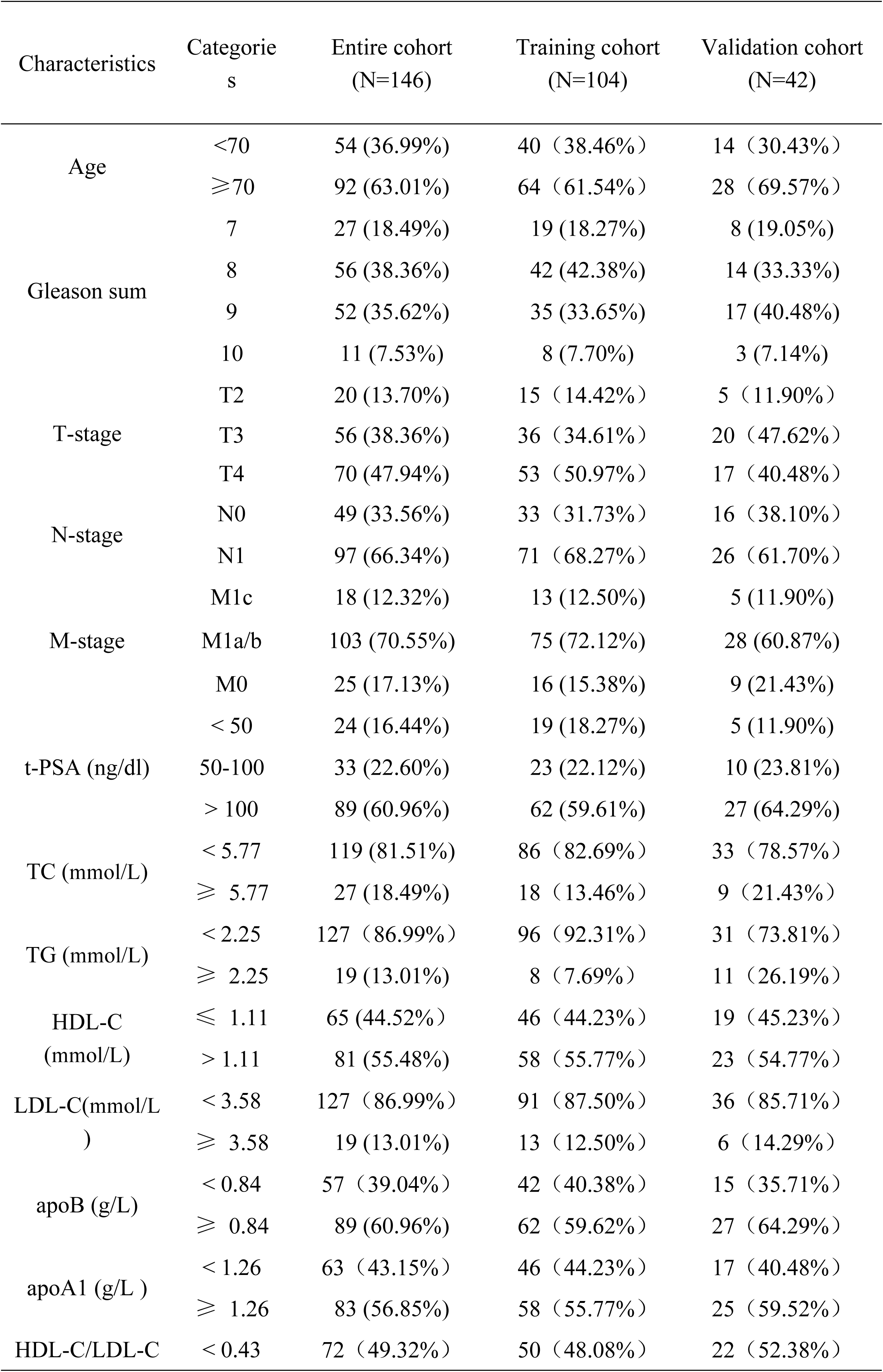
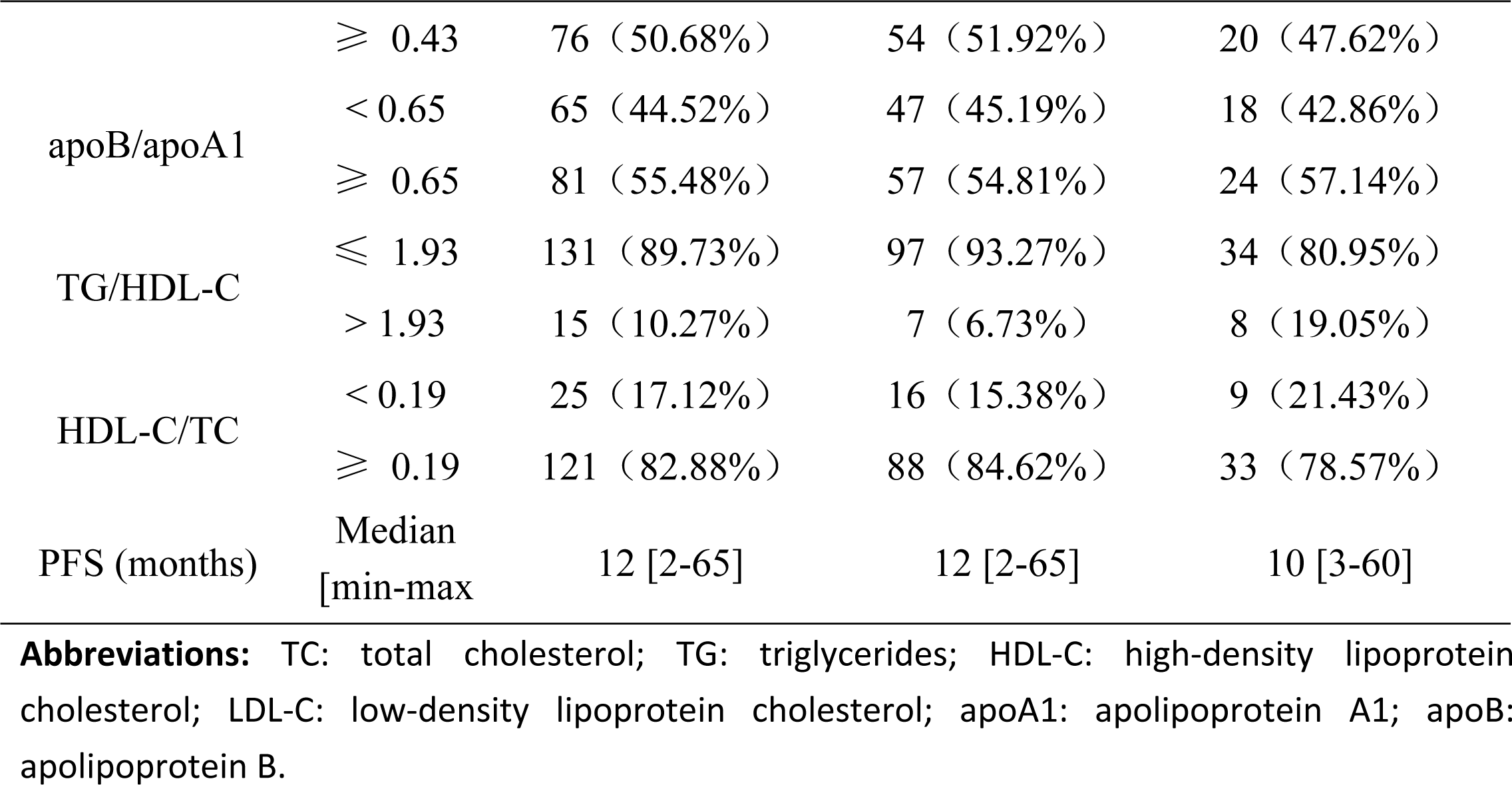
Representativeness of study participants

### Establish a nomogram for predicting the CRPC progression

In this study, 146 patients were randomly assigned to training set of 102 samples (∼7/10), and internal validation set of 44 samples (∼3/10). The R package “survival” was used to integrate data on survival time, survival status, and the clinico-pathological characteristics. The multivariate Cox regression analysis was used to determine the prognostic significance of these characteristics in the training set. On the basis of multivariate Cox proportional hazards analysis, nomographs predicting rates of disease progression at 6th, 12th, 18th, and 24th months were created using the “RMS” software. The nomogram presents graphical data for these factors, and from the points linked with each risk factor, the prognosis risk of an individual patient may be computed. During the validation of the nomogram, the total points of each patient in the validation cohort were calculated according to the established nomogram. The concordance index (C-index), calibration curves, receiver operating characteristic (ROC) curves, and decision curve analyses (DCA) were used to evaluate the clinical utility of nomogram.

### Analyzed the survival patients with high-risk or low-risk score based on nomogram

The best risk score cut-off value was determined for the entire dataset using the R software program maxstat (Maximally selected rank statistics with multiple p-value approximations, version: 0.7-25). The minimum and maximum numbers of samples in each category were set at more than 25% and less than 75%, respectively. Based on available information, high-risk and low-risk groups of patients were separated. The Survfit function of the R software program was used to investigate the prognostic disparity between the two groups, and the significance of the prognostic difference between the several groups of samples was assessed using the log-rank test.

### Statistical analysis

The ideal cutoff values for age, lipid index and their derivatives were determined using X-tile software v3.6.1 and all statistical analyses were performed using SPSS version 24.0. For categorical variables, frequencies and proportions are provided. Using multivariate Cox regression analysis, the relevant hazard ratios (HRs) and 95% confidence intervals (CIs) were calculated. For DCA, the “ggDCA” R package was utilized. The area under the curve (AUC) was calculated by ROC analysis using the R software package pROC (version 1.17.0.1). We specifically collected the patients’ follow-up duration and risk score, then ROC analysis were performed using the ROC function of pROC at 6, 12, 18, and 24 months. For all analyses, a P value of < 0.05 was considered statistically significant.

## Results

### Baseline clinicopathological characteristics

A total of 146 patients who fulfilled the criteria for lipid and survival information were enrolled and were separated into the training (n=102) and validating (n=44) cohorts. Overall, the median age at diagnosis was 74 (50, 92) years. At the time of diagnosis, 18 (12.32%), 103 (70.55%) and 25(17.13%) patients were diagnosed as M1c, M1a/b and M0 stage. 97 (66.34%) patients had lymph node metastasis. 70 (47.94%), 56 (38.36%) and 20 (13.70%) patients were diagnosed as T4, T3 and T2 stage, respectively. Among the Gleason sum values, 27 (18.49%), 56 (38.36%), 52 (35.62%) and 11 (7.53%) had a score of 7, 8, 9 and 10, respectively. The median PFS was 12 months, ranged from 2 to 65 months. We divided normal distribution continuous variables, including the age, lipid index and their derivatives, into categorical variables based on the optimum cut-off values. Detailed characteristics in the cohorts were summarized in Table 1.

### Construction of a combined Nomogram for individualized prediction

The clinico-pathologic factors, including age, PSA levels, clinical TNM stage, Gleason sum, TC, TG, HDL-C, LDL-C, apoA1, apoB and HDL-C/LDL-C, TG/HDL-C, apoB/apoA1, HDL-C/TC were used for univariate and multivariate Cox regression analyses (Table 2). The results demonstrated that TC, LDL-C, apoB, N stage and Gleason sum were independent risk factors for HSPC progression (Table 2). By the multivariate Cox regression analysis, a nomogram integrated with the TC, LDL-C, apoB, N stage and Gleason sum was established in the training cohort (Figure 1). The C-index was 0.740, (95% CI, 0.662-0.818), indicating a good consistency. The calibration plots (Figure 2A) showed that the nomogram performed well in the individualized prediction of progression to CRPC. DCA curve revealed that the nomogram provided obvious net benefit at 6th, 12th, 18th and 24th month to the none or all strategy, which is better than that of NG model (N stage plus Gleason sum) within the actual threshold probability range (Figure 2B). As shown in the ROC curve, the 6th, 12th, 18th and 24th AUC of the nomogram were 0.896, 0.765, 0.793 and 0.794, respectively (Figures 2C), all of which were superior to the AUC values predicted by the NG model.

**Figure 1.**
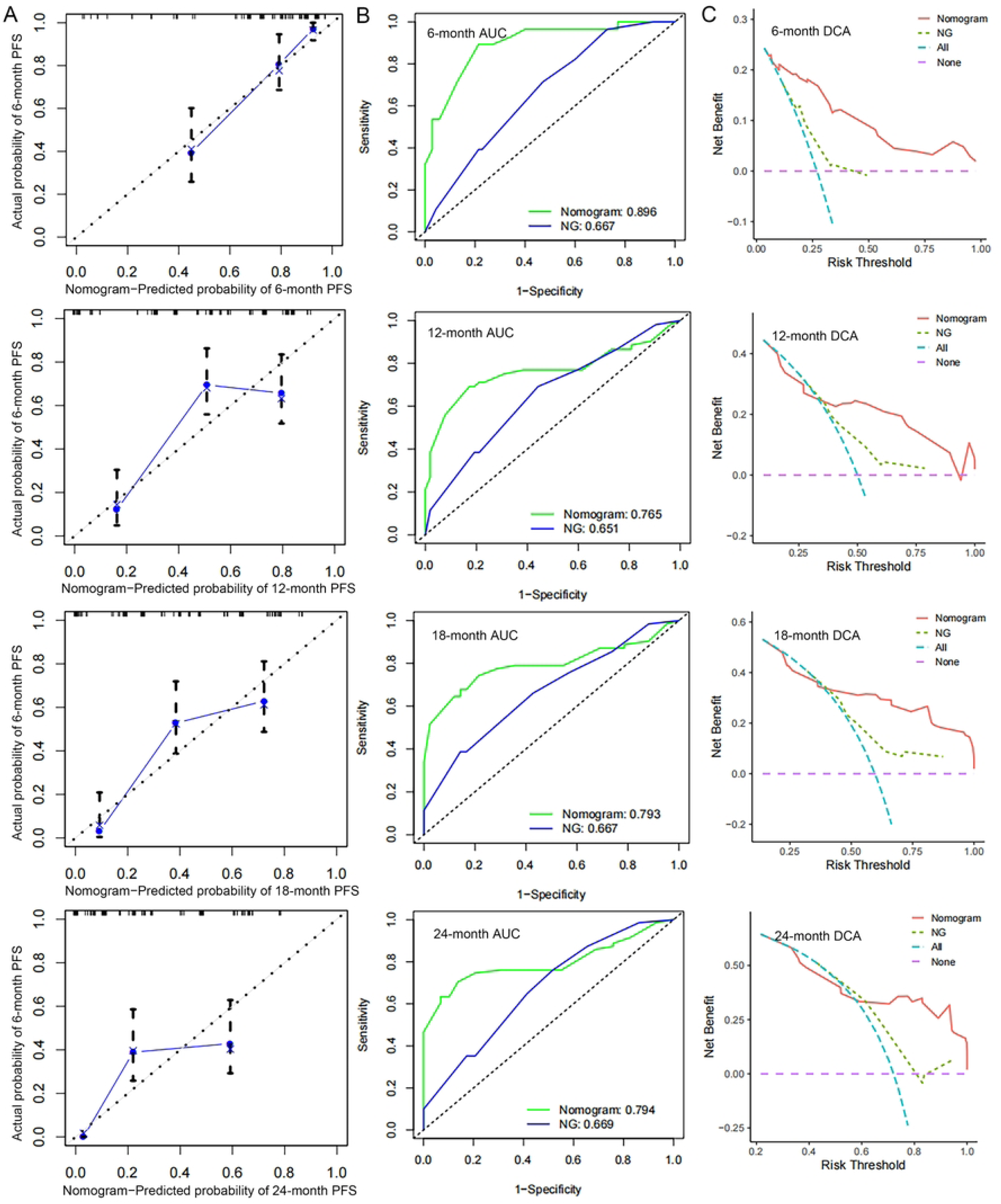
Establishment and validation of a combined nomogram in the training cohort. (A) Nomogram based on total cholesterol (TC), apolipoprotein B (apoB), low-density lipoprotein cholesterol (LDL-C), N stage and Gleason sum was constructed to predict the 6-, 12-, 18- and 24-month CRPC-free survival.

**Figure 2.**
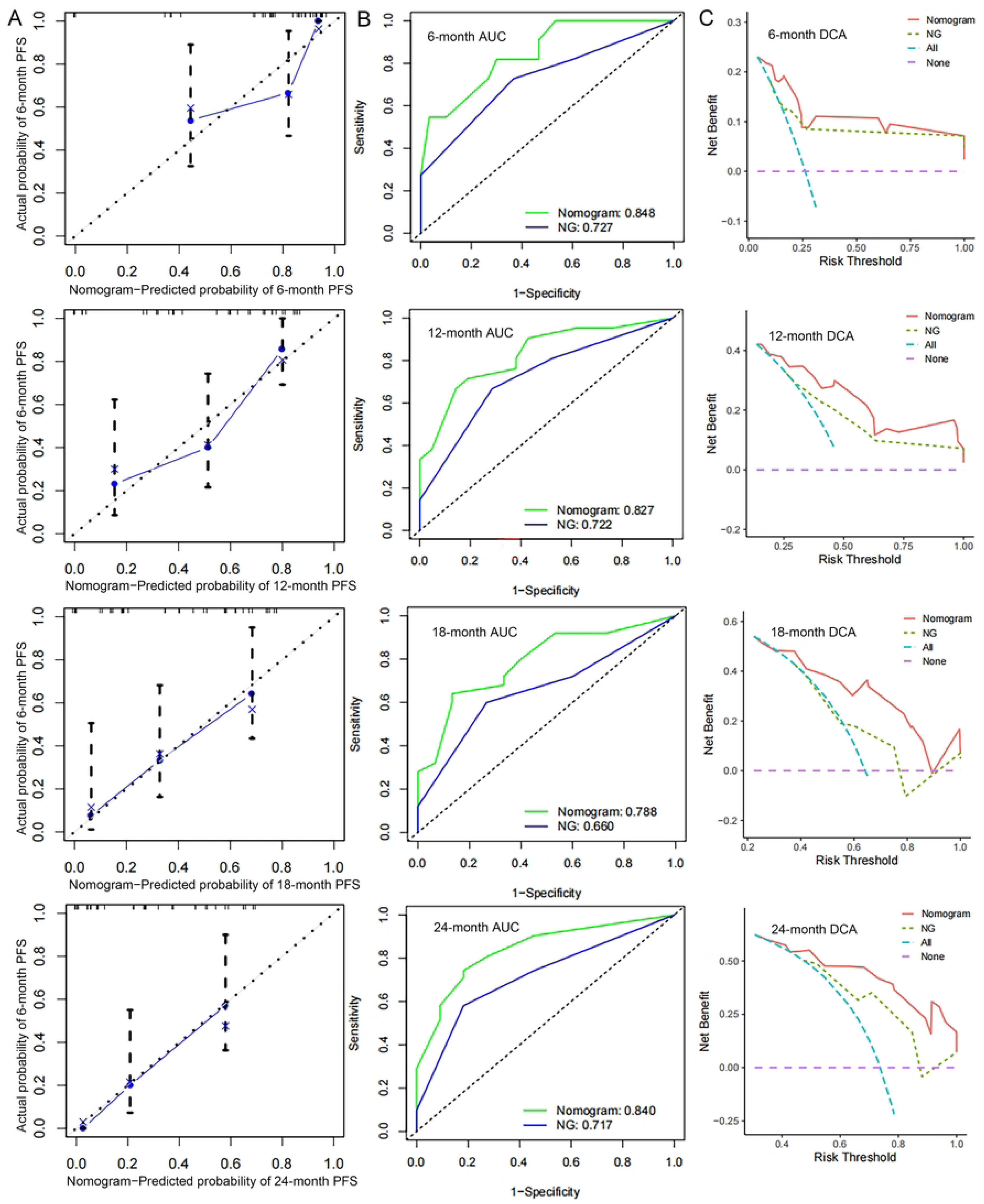
Validation of predictive capacity of the Nomogram in the training cohort. (A) Predictive accuracy of the nomogram was assessed by the calibration plots; (B) Comparing ROC curves of the nomogram and NG model (N stage plus Gleason sum) for 6-, 12-, 18- and 24-month CRPC-free survival; (C) Comparing the time-dependent decision curve analysis for the clinical benefit of the nomogram and NG model.

**Table 2.**
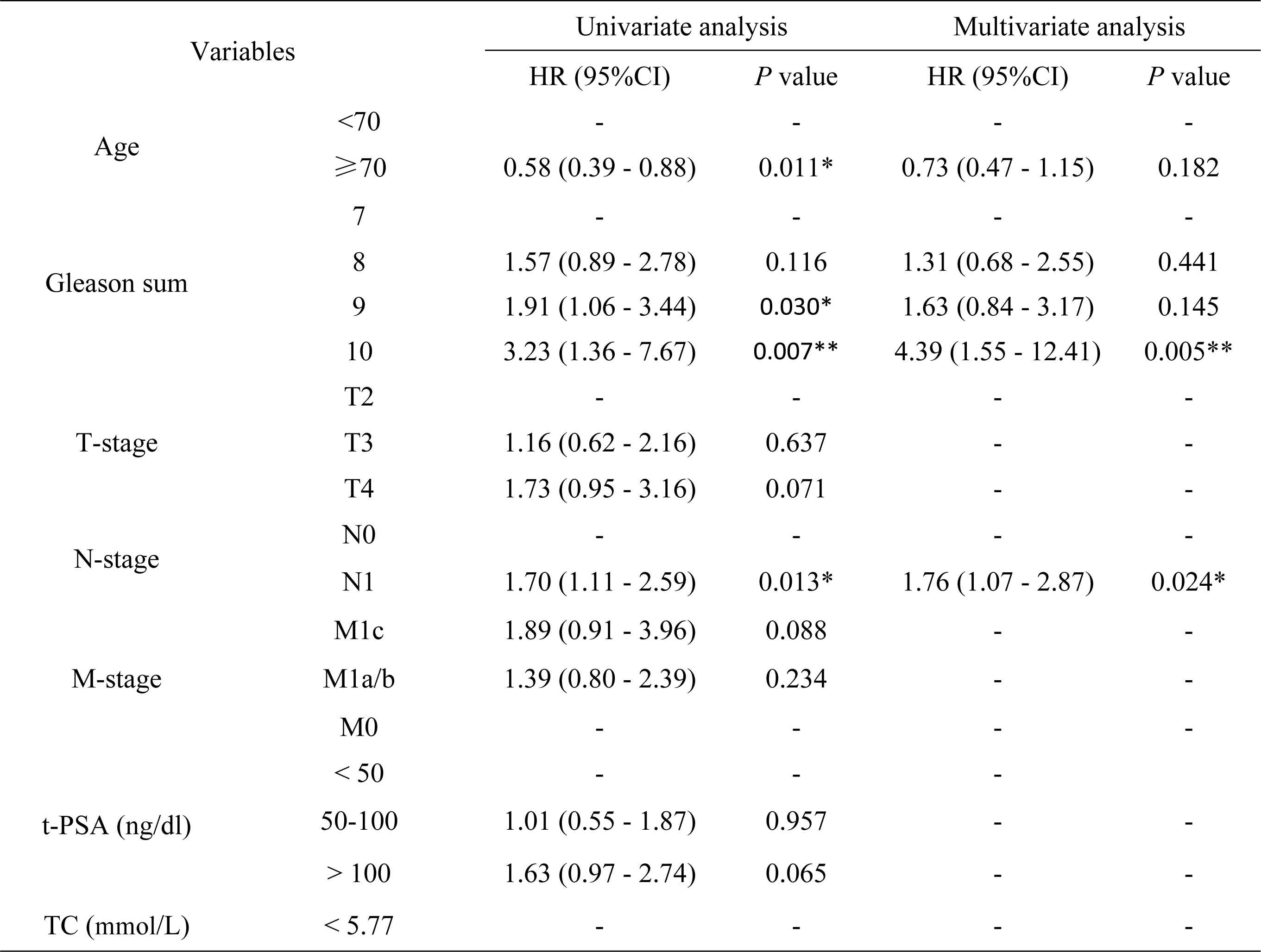
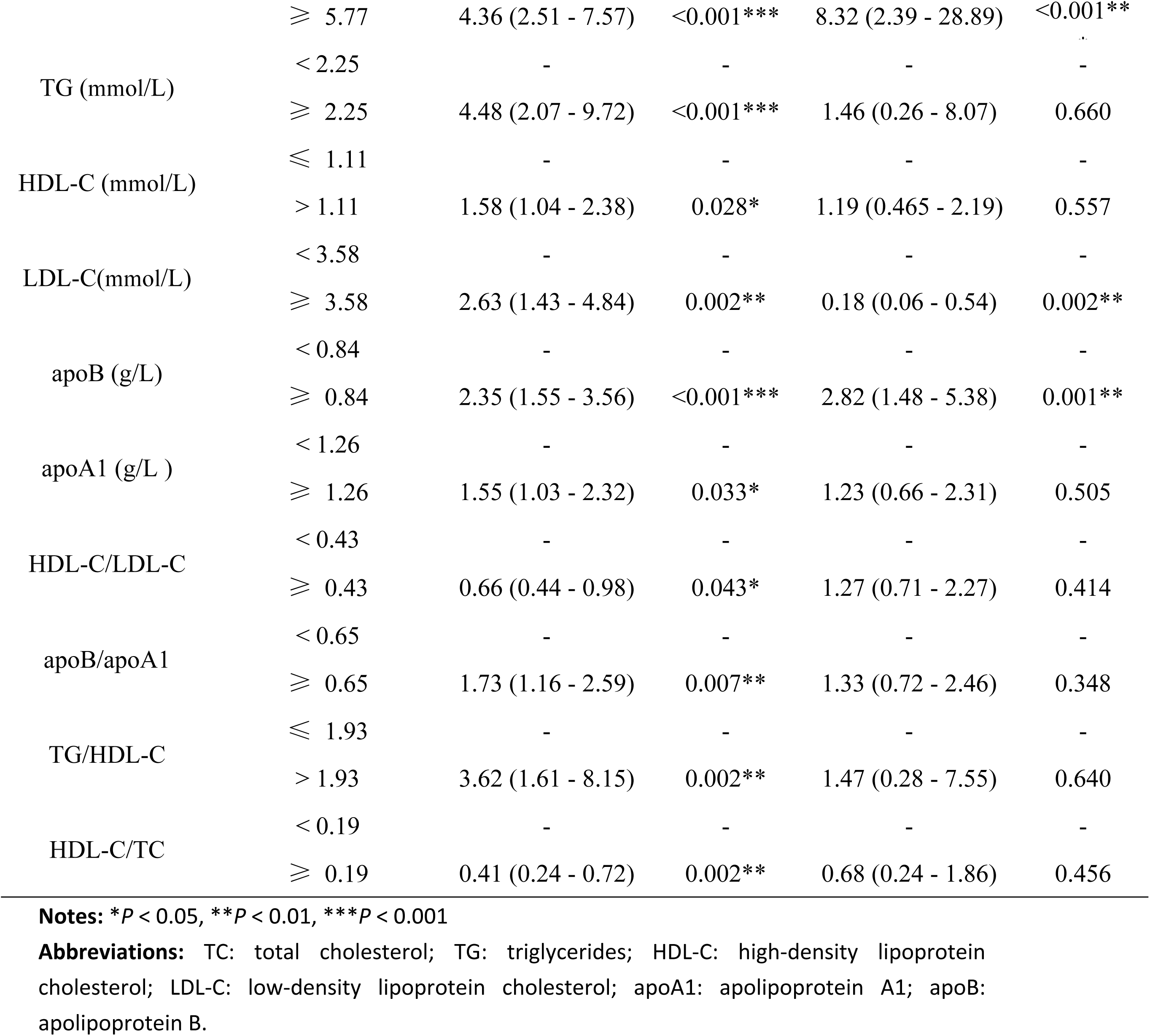
Univariate and multivariate analyses of factors associated with HSPC progression

### Predictive accuracy validation of the Nomogram

In the validation cohort, the C-index of the nomogram for predicting CRPC progression was 0.755 (95% CI, 0.677-0.814), the calibration curve showed good agreement between prediction and observation in the probability of 6th, 12th, 18th and 24th survival (Figure 3A). DCA curve also demonstrated positive net benefits in guiding clinical decisions (Figure 3B). Anymore, the 6th, 12th, 18th and 24th AUC were 0.848, 0.827, 0.788 and 0.840, respectively, (Figure 3C), all of which were superior to the AUC values predicted by the NG model. Through nomogram modeling, all patients in this study were turned into a single risk score. Furthermore, the best risk score cutoff value was calculated. The minimum sample size was set at greater than 25 percent, while the maximum sample size was set at less than 75 percent. Based on this information, patients were split into high and low risk groups, with 46.6 percent (n = 68) in the high risk group and 53.4 percent (n = 78) in the low risk group. Between the two groups, there was a statistically significant difference in prognosis (Figure 4).

**Figure 3.**
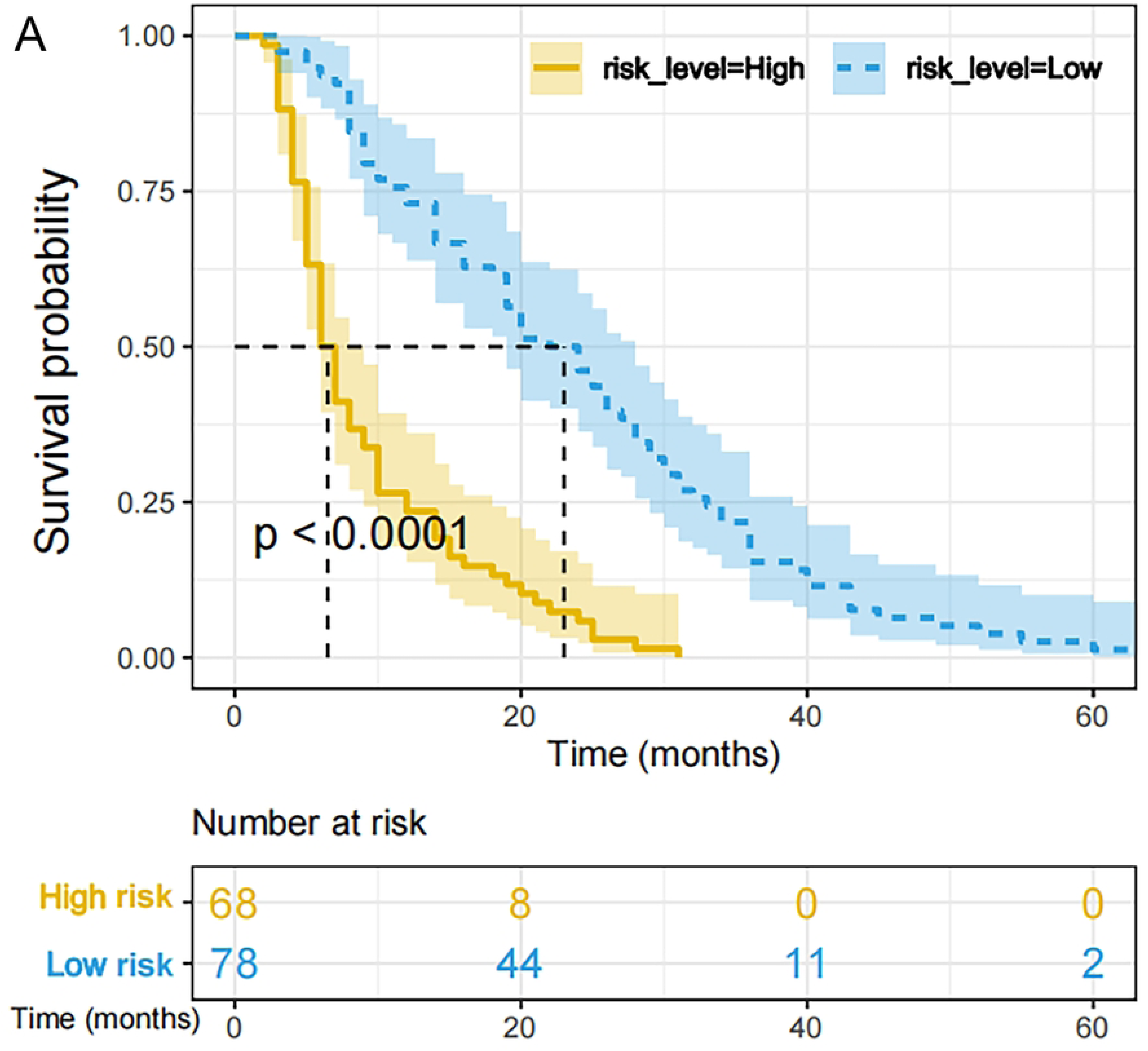
Validation of predictive capacity of the Nomogram in the testing cohort. (A) Predictive accuracy of the nomogram was assessed by the calibration plots. (B) Comparing ROC curves of the nomogram and NG (N stage plus Gleason sum) model for 6-, 12-, 18- and 24-month CRPC-free survival. (C) Comparing the time-dependent decision curve analysis for the clinical benefit of the nomogram and NG model.

**Figure 4.**
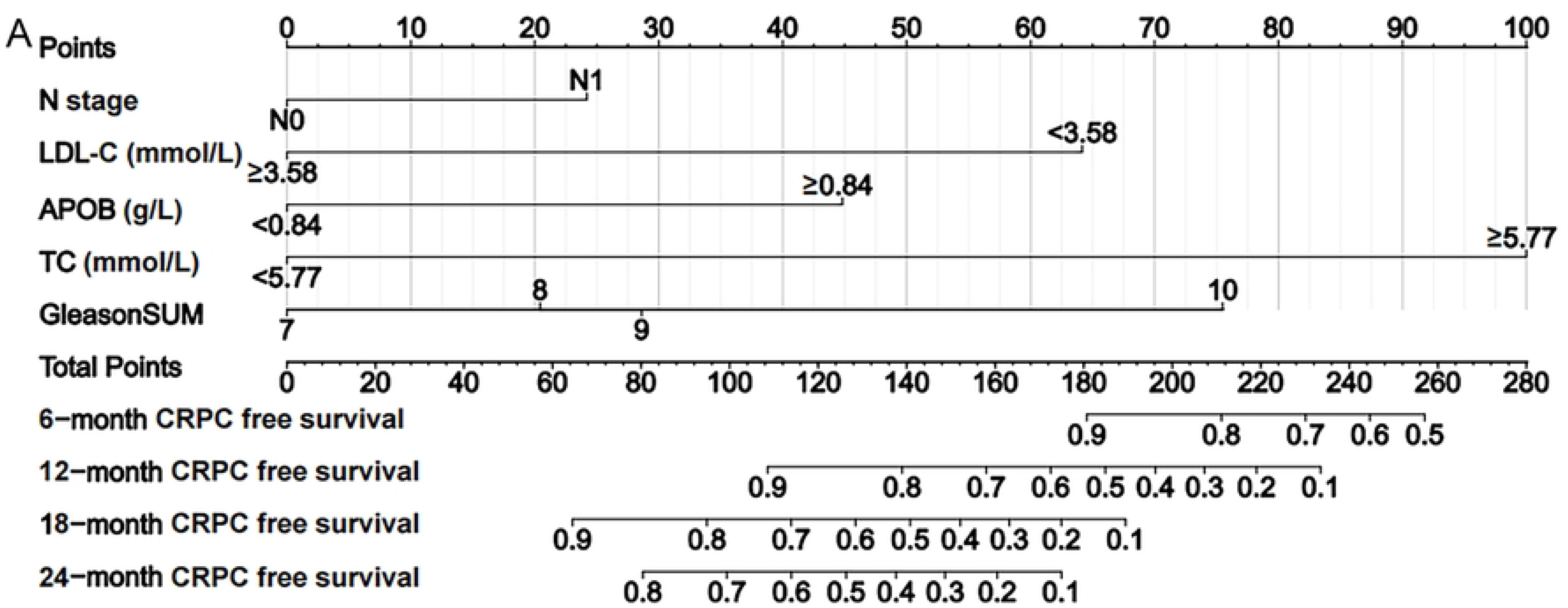
Validation of predictive value of the Nomogram. (A) The progression free survival curves based on nomogram correlated risk score in the whole cohort.

## Discussion

In this study, we focused on the relationship between serum lipid and clinical outcome in advanced HSPC patients, who received LHRH agonists coupled with anti-androgen as initial and only therapy before progression. Through multivariate cox analysis, Furthermore, we established a nomogram model containing TC, LDL-C, apoB, N stage and Gleason sum, hypothesized that its predictive significance would help clinicians to identify high-risk HSPC patients in a timely manner as well as to provide reasonable treatment options.

Disruption of lipid metabolism in cancer cells can cause structural alterations to the tumor cell membrane, turbulence in intracellular energy metabolism, and dysregulation of cell signal transduction and gene expression [13]. As a source and transforming component of lipid metabolism in the tumor microenvironment, serum lipids play an important role in carcinogenesis [14]. There has, however, been no extensive study of the link between serum lipids and advanced HSPC.

Using multiple regression analysis, we concluded that there lipid index, TC, LDL-C and apoB were independent risk factors for rapid progression of HSPC patients. Cholesterol is an important component of the cell membrane and is required for the integrity of the lipid rafts, which stimulates cell proliferation [15]. Patients with colorectal cancer and cervical cancer who have elevated total serum cholesterol levels have a poor prognosis, according to the research [16, 17]. Our research consistently demonstrated that greater serum TC levels were an independent risk factor for HSPC development. Patients with gastric cancer and primary liver cancer who have decreased total serum TC levels have a worse prognosis [18, 19]. The notion was that rapid proliferation of cancer cells, particularly in the late stage, necessitates the participation of significant amounts of cholesterol, promoting total cholesterol depletion [20].

LDL-C is a component of cholesterol that participates in cholesterol transport. According to Saito et al., LDL-C levels are connected with an increased risk of developing hepatocellular carcinoma [21]. LDL-C levels were found to be a negative predictor of disease-free survival in breast cancer patients by Rodrigues et al. [22]. Our study also found that high-level LDL-C in HSPC patients were associated with a poor prognosis. The contradictory assessment of the prognostic role of LDL-C may be explained by several hypothesis. Benn et al. reported that although low plasma LDL-C correlated with an increased risk of cancer, this was not the case for a patient with genetically decreased LDL-C [23].This suggests that low LDL-C concentrations may not cause cancer in and of themselves, but are more likely to be caused by associated nutritional deficiencies that emerge when cancer advances. The LDL receptor, on the other hand, has been reported to be overexpressed in several cancers, boosting LDL-C absorption and new membrane synthesis to fulfill the need of cancer cells [24]. Furthermore, increasing reactive oxygen species levels during an inflammatory state might cause LDL to be oxidized to oxidized LDL (ox-LDL). This causes a drop in circulating LDL because ox-LDL is taken up by macrophages at the site of inflammation [25].

The apoB is is the primary apolipoprotein of chylomicrons, VLDL, IDL, and LDL particles, and it is responsible for delivering fat molecules to all peripheral tissues as well as having a crucial role in carcinogenesis [26], which may help to explain the link between high apoB levels and poor HSPC prognosis. The epidemiological evidence for the link between apoB levels and cancer prognosis is debatable. Borgquist S, et al. observed in a sample of 28,098 patients that increased apoB was related with cancer risk in colorectal cancer, lung cancer, and breast cancer [26]. Sun et al. discovered that high levels of apoB might predict a poor outcome in malignant biliary tumors [27]. In contrast, Chen et al [28] found that high levels of apoB were helpful for the prognosis of individuals with lung cancer in a research on tumor prognosis. The inconsistent results in the aforementioned studies could be attributed to differences in the study population, follow-up length, outcomes, and statistical adjustment for confounding factors. Furthermore, earlier research used different blood lipid cut-off values, and these investigations did not exclude patients who were using cholesterol-lowering medicines.

Above all, with the aid of multivariate Cox regression analysis, a novel HSPC prognostic nomogram integrating TC, LDL-C, apoB, N stage and Gleason sum was established, and suggested more favorable discriminative and predictive ability compared with the N stage and Gleason sum, which were identified by ROC curve and DCA analysis. After dividing the patients into groups with low- and high-risk score, the group with a high-risk score developed rapidly and had a bad prognosis. Thus, the nomogram based risk system provide a convenient and intuitive tool to initially classify advance HSPC patients into different prognostic stage.

This research is a single-center study with a small sample size, more multi-center, large-sample investigations are needed to confirm the association between serum lipids and HSPC clinical characteristics and prognosis. We will perform additional molecular research on the interaction between lipid metabolism and the biological behaviors of PCa cells, closely linking them to patients’ serum lipid levels and prognosis, bringing new ideas to the therapy of HSPC.

The successful construction of a novel nomogram for HSPC patients based on serum lipid profiles has revealed fresh knowledge about the association between serum lipids and HSPC progression. This knowledge will assist clinicians in quickly and accurately assessing the prognosis of patients and identifying high-risk HSPC in order to develop individualized treatment plans and follow-up strategies.

## Statements and Declarations

### Disclosure

**-Conflict of interest :** The authors declare that they have no known competing financial interests or personal relationships that could have appeared to influence the work reported in this paper.

**-Ethics approval:** The research protocol was accepted by the ethical committee of the Second Hospital of Dalian Medical University (number: 2023064) with the Declaration of Helsinki, .

-**Registry and the Registration No. of the study/trial :** N/A

**-Informed Consent:** all patients submitted written informed consent prior to analysis

**-Animal Studies :** N/A.

## Data Availability

The data analyzed during the current study are available from the corresponding author on reasonable request.

## Acknowledgement

This work received technical supported from the Second Affiliated Hospital of Dalian Medical University, Dalian, Liaoning, China.

## Author contributions

M.W. : Methodology, Software, Data Curation, Writing - Original Draft. Y.H. : Methodology, Software, Data Curation, Writing - Original Draft. C.P..: Writing - Review & Editing, Supervision, Validation. Y.Z. : Methodology, Software, Data Curation, Writing - Original Draft. B.Y.: Writing - Review & Editing, Supervision, Project administration, Funding acquisition.

